# Scalable screening for emergency department missed opportunities for diagnosis using sequential eTriggers and large language models

**DOI:** 10.1101/2025.10.06.25337201

**Authors:** Clifford Marks, Sean Gibney, Bryan Stenson, Deesha Sarma, Cynthia Gaudet, Haadi Mombini, Thomas Buckley, Laura Burke, Nathan I. Shapiro, Jonathan L. Burstein, Shamai A Grossman, Anika Parab, Alexander T. Janke, Arjun Manrai, Richard Andrew Taylor, Carlo L. Rosen, Adam Rodman, Adrian D. Haimovich

**Affiliations:** Department of Emergency Medicine, MedStar Georgetown University Hospital, Georgetown University School of Medicine, Washington, DC; Department of Emergency Medicine, Beth Israel Deaconess Medical Center, Harvard Medical School, Boston, MA; Department of Emergency Medicine, Boston Medical Center, Boston University Chobanian & Avedisian School of Medicine, Boston, MA; Beth Israel Lahey Health Technology and Innovation, Beth Israel Lahey Health, Boston, MA; Department of Biomedical Informatics, Harvard Medical School, Boston, MA; Department of Emergency Medicine, Warren Alpert Medical School of Brown University, Providence, RI; Department of Emergency Medicine, Lahey Hospital & Medical Center, UMass Chan-Lahey School of Medicine, Burlington, MA; Department of Emergency Medicine, University of Michigan Medical School, Ann Arbor, MI; Department of Emergency Medicine, University of Virginia School of Medicine, Charlottesville, VA; Department of Medicine, Division of General Medicine, Beth Israel Deaconess Medical Center, Harvard Medical School, Boston, MA

## Abstract

**Importance:** Missed opportunities for diagnosis (MODs), sometimes termed diagnostic errors, are a major cause of patient morbidity and mortality in the emergency department (ED). EDs have employed eTriggers, rule-based case collections likely to have a higher than average error rate (e.g. 72 hour returns with admission), but their utility is limited by low error yields. Large language models (LLMs) offer new opportunities to identify MODs and contribute to both individual- and systems-level quality improvement.

**Objective:** To determine whether sequential screening of ED cases with eTriggers and an LLM can more efficiently identify MODs compared to eTriggers alone.

**Design:** Retrospective observational cohort study of ED encounters collected between March 2015 and June 2025.

**Setting:** 10 EDs (2 academic, 8 community) in a single US health system.

**Participants:** Emergency physicians reviewed and adjudicated random samples of cases identified by 3 previously validated eTriggers (72-hour return with admission, 10-day return with ICU admission, and floor-to-ICU escalation within 24 hours) using the SaferDX instrument. An ED physician also evaluated a novel hybrid eTrigger combining an LLM adjudicator with a rules engine for 9-day return admissions with emergency care– sensitive conditions (ECSCs).

**Exposures:** LLM MOD adjudication of ED cases with Claude Sonnet 4 using an iteratively-developed, standardized prompt incorporating the SaferDx instrument.

**Main Outcome(s) and Measure(s):** Positive predictive value (PPV), sensitivity, specificity, negative predictive value (NPV), and number needed to screen (NNS) for MODs. Reviewer time to adjudicate cases and quality improvement stakeholder assessments of LLM case summaries were also measured.

**Results:** Of the 357 encounters (mean [SD] age, 65.2 [17.8] years; 47.1% female) reviewed, adjudicated MOD PPV ranged from 11.0% to 18.6% across traditional eTriggers. For 72-hour return admissions, the LLM achieved sensitivity 85.7% (95% CI, 65.4%-95.0%), specificity 56.8% (95% CI, 49.3%-64.0%), PPV 19.8%, and NPV 97.0%. For 10-day ICU returns, sensitivity was 100% (95% CI, 56.6%-100%), specificity 43.5% (95% CI, 25.6%-63.2%), PPV 27.8%, and NPV 100%. For floor-to-ICU escalations, sensitivity was 55.6% (95% CI, 33.7%-75.4%), specificity 64.6% (95% CI, 53.6%-74.2%), PPV 26.3%, and NPV 86.4%. The hybrid ECSC eTrigger identified 110 MODs (53.1% of 207 encounters), with blinded review of a stratified sample estimating PPV 45% and NPV 100%. Expert reviewers required a median of 5 minutes per case; restricting review to LLM-positive charts reduced review time by up to 50% without missed errors for these triggers. In stakeholder review, LLM-generated case summaries were rated highly actionable for individual clinician feedback (mean, 4.1 of 5) but less so for systems-level interventions (mean, 1.4 of 5).

**Conclusions and Relevance:** In this multisite retrospective study, LLMs demonstrated high NPVs across multiple eTrigger criteria. Sequential use of LLM and human review improved efficiency and detection compared with traditional eTriggers, and narrative case summaries offered a novel method to identify opportunities for clinician-level feedback. These findings suggest that LLM-based approaches may provide scalable diagnostic quality oversight in the ED.

**Key Points:** *Question:* Can sequential screening with eTriggers and a large-language-model (LLM) identify missed opportunities for diagnosis (MODs) in the emergency department, improving screening efficiency versus traditional eTriggers?

*Findings:* In a multicenter retrospective cohort (10 EDs; 317 reviewed encounters), LLM adjudication showed high sensitivity and NPV across three established eTriggers (e.g., 72-hour returns: sensitivity 85.7%, NPV 97.0; 10-day ICU returns: sensitivity 100%, NPV 100%). A sequential approach was validated on a novel eTrigger for 9-day returns for select emergency care sensitive conditions, achieving PPV 45% and NPV 100% in 40 blinded samples.

*Meaning:* LLM-augmented eTrigger screening offers scalable, efficient MOD detection to support diagnostic quality oversight in EDs.

## Introduction

Missed opportunities for diagnosis (MODs), also known as diagnostic errors, are a leading cause of iatrogenic harm, potentially accounting for tens of thousands of deaths annually in the United States, and despite decades of mitigation efforts, they remain an intractable problem.^1,2^ This stems in part from difficulties associated with defining and measuring MODs themselves. Process measures of MODs rely on robust investments in expert labor, with time-consuming and costly manual chart review.^3^ Even then, MODs rarely have an absolute truth, and expert reviewers often demonstrate significant levels of disagreement.^4^ Researchers have increasingly focused on eTriggers as an initial screen for MOD review - i.e., cases identified based on administrative data as particularly high risk for error.^3,5–7^ Confining review to cases high-risk for MODs better uses finite resources, but these limited case sets still have low yields for diagnostic error.^5,6,8^ Disease-specific pathways to assess MODs suffer from similar drawbacks, since these omit large numbers of diagnostic errors and may obscure contributing systems-level processes across disease entities.^5,9^

Large language models (LLMs) capable of rapidly analyzing large quantities of clinical information and summarizing complex episodes of care without extensive human labeling,^10^ are a promising tool for error identification. A recent study of a commercial LLM (ChatGPT, OpenAI) applied to a case series of previously identified MODs showed that the model was very sensitive in identifying potential errors and also could assign more contributing factors than expert reviewers.^11^ Expert-like performance has been observed in radiology report error identification.^12,13^ Model performance continues to improve with so-called “reasoning models,” which use run-time inference and chain-of-thought to improve their reasoning abilities.^14^ Recent research on human and LLM assessments of both clinical vignettes and ED cases shows LLMs outperforming human clinicians at both generating differential diagnoses and developing diagnostic and management plans.^14^ Despite these promises, LLMs have unique drawbacks in MOD adjudication, including biased reasoning and a propensity to hallucinate i.e., producing inaccurate information due to their generative nature.^14,15^ Furthermore, LLMs are subject to the same taxonomic uncertainties that affect humans in adjudicating MODs, and their test characteristics in a real world setting remain unknown.

The emergency department (ED) represents a particularly important environment for addressing MODs owing to unique factors that may both make such errors potentially more prevalent and more consequential than in other settings.^16–18^ In this study, we examined the ability of an LLM to assess ED cases for MODs by comparing their conclusions to a gold standard of two-physician agreement across three sets of eTrigger cases. We then assessed the generalizability of this sequential screening approach by testing the MOD performance of a novel sequential ED eTrigger integrated with an LLM.^19^ Finally, we assessed the potential value of brief case narratives generated by the LLM for informing future diagnostic error prevention.

## Methods

### Study design and setting

This was a retrospective observational cohort study of encounters from 10 hospitals (2 academic, 8 community) across the Beth Israel Lahey Health enterprise health system. Study dates were between March 2015 and June 2025. This study was approved by the BIDMC IRB (2025P000212). No patients or public members were involved in study design or conduct as this was a retrospective analysis of existing clinical data. We followed the TRIPOD-LLM guidelines (Supplemental Materials).^20^

### eTriggers implementation

eTriggers are administrative criteria to identify cases at high risk for MODs. Based on prior literature and the clinical experience of the emergency physicians involved, we implemented two discharge eTriggers and one admission eTrigger (Appendix A).^5^ The discharge triggers were ED discharge with return and admission within 3 days (72-hour return to admission) and ED discharge with return to direct ICU admission in the next 10 days (10-day return to ICU admission).^21^ The 3 day return timeline was selected to align with current organizational review criteria and prior ED quality literature.^22^ Where multiple ED visits preceded the return admission, only the most recent ED discharge before the admission visit was used. The admission eTrigger was non-ICU admission with an upgrade to ICU level of care within 24 hours (floor-to-ICU).^6^ For 72 hour returns, a stratified sample was used: the first set comprised a random selection of cases from an academic ED from June 2024 to February 2025, while the second set comprised a random selection of cases from across the hospital system from March 2015 to March 2025. All other eTriggers sampled cases from across the hospital system from January 2024 to June 2025. Custom SQL queries were developed using inclusion criteria described in Appendix A using our enterprise Snowflake data warehouse.

### Physician adjudication of missed opportunity for diagnosis for previously described eTriggers

In alignment with prior literature, we randomly sampled cases from each of the three previously described eTrigger scenarios (72-hour return to admission, 10-day return to ICU admission, and floor-to-ICU). All 317 sampled eTrigger cases (100%) were reviewed by two emergency physicians using the following question: was there a missed opportunity to make a correct or timely diagnosis based on the available evidence, regardless of harm? Reviewers were emergency physicians representing 3 different hospitals with a median 7 years of practice experience. Each reviewer was provided with specific instructions that included the SaferDx Instrument (Appendix B). To ensure rubric alignment, a training subset was reviewed by all case reviewers. Physician coders were asked to adjudicate disagreements to establish a gold standard for each case and persistent disagreements were resolved by a third reviewer. Reviewers also flagged cases that did not appear to meet criteria for inclusion (e.g. miscoded as ICU upgrade, no associated ED note), and these were excluded if determined to not meet inclusion criteria by the study authors. Reviewers were asked to time themselves doing a subset of cases and the median and interquartile range for per-case review is presented. These adjudicated cases were used to present the positive predictive value (PPV), number-needed-to-screen (NNS) to identify one error, and time to identify one error for each of the eTriggers. Google Gemini and OpenAI ChatGPT were used to develop SQL and Python code (v 3.11.8) for analysis.

### Comparison to language model MOD determination

Prompt development followed a structured iterative process: (1) Initial prompts were adapted from human reviewer instructions, incorporating the SaferDx instrument; (2) Two physicians (ADH, CM) tested prompts on 5-10 cases per eTrigger category, comparing LLM outputs to their own clinical judgment; (3) Prompts were refined based on failure modes - for example, adding ‘You are NOT to consider management, only diagnosis’ after the LLM initially conflated treatment and diagnostic errors; (4) Final prompt selection required agreement between authors that LLM reasoning aligned with emergency physician diagnostic logic. The final prompts (Appendices C-E) were then frozen before full-scale analysis. As we employed a pre-trained commercial LLM (Claude Sonnet 4) without additional training or fine-tuning, there was no development dataset requiring comparison to evaluation data. The same diagnostic error prompt was used across the three but incorporated different clinical data depending on the eTrigger (Appendix D). The LLM was asked to return structured JSON output containing: (1) a binary MOD classification (’Yes’/’No’), (2) a probability estimate (0-100%), and (3) an unstructured text rationale with diagnostic reasoning. For primary analyses, we used the binary classification directly without post-processing. The narrative rationales were used without modification for the stakeholder review sessions (Table 3) to assess actionability for quality improvement.

We used the Claude Sonnet 4 model (20250514-v1:0, with training data through January 2025) accessed via an enterprise-managed, PHI-compliant Amazon Web Services entrypoint. LLM analysis was performed between April 2025 and June 2025. The maximum number of tokens used was 4096 and the temperature was 0.1. LLM predictions were obtained via API calls to Claude Sonnet 4, with the binary ‘mod_decision’ field extracted from the JSON response. Processing each encounter took approximately 30 seconds. Performance was assessed against the gold standard (two-physician agreement with third-reviewer adjudication) using standard diagnostic test metrics: sensitivity, specificity, positive predictive value (PPV), and negative predictive value (NPV).

### Development and validation of a novel, LLM-integrated eTrigger for 9-day high-risk emergency care sensitive conditions

We implemented a hybrid eTrigger comprising (1) a rules engine identifying ED discharges with return admission within 9 days for one of ten emergency care–sensitive conditions (ECSCs), based on recent Medicare claims benchmarking,^19^ and (2) an LLM adjudicator using a standardized missed-opportunity (MOD) prompt (Appendix A, E). Query dates were January 2024-January 2025. We present MOD rates stratified by ECSC. A random sample of 20 LLM-positive and 20 LLM-negative cases were reviewed by a blinded reviewer who was given the mixed, shuffled sample to estimate PPV and NPV with 95% Wilson score CIs. For each of the ten ECSCs studied, we then report the percent of cases identified by the LLM as having an MOD.

### Quality improvement stakeholder case review

Two emergency physician authors (ADH, CM) extracted a subset of cases of LLM-identified MODs from across all four cases described (72-hour return to admission, ED discharge with ICU return admission within 10 days, floor-to-ICU escalations, and ECSC within 9 days of ED discharge) and presented only the AI-generated narrative with the suspected missed diagnosis to a panel of emergency medicine quality and safety experts drawn from multiple sites across the enterprise health system. For each case, the panel was first asked to discuss the case and insights for both systems-level and individual-level interventions. After discussion, a moderator instructed the panel to reach a consensus on three questions, each rated on a 5-point Likert scale: Q1. Would you recommend this case for further chart review by a member of the QA team? (responses: definitely would not recommend to definitely recommend); Q2. From a systems quality perspective, is the case described by this summary potentially actionable? (responses: this case would definitely not change system quality performance to it definitely would); Q3. From an individual clinician quality perspective, is the case described by this summary potentially actionable? (responses: this case would definitely not change system quality performance to it definitely would). Consensus Likert responses for each question were recorded.

## Results

Individual eTrigger demographics from reviewed cases are included in Table 1. In our sample, there were 110,281 admissions of which 281 cases met floor-to-ICU inclusion criteria and 100 were randomly sampled for review (Figure 1A). Three were subsequently excluded - two were direct ICU admissions that were miscoded and the third went to the operating room from the ED and then to a surgical ICU. After adjudication, there were 18 MODs for a PPV of 18.6% and a NNS of 5.4. Of 1,790 ED admissions to the ICU in our sample, there were 30 cases with an ED discharge within the preceding 10 days of which one was excluded as the patient chart was incomplete. Of these there were 5 MODs for a PPV of 17.2% and NNS of 5.8. In the academic ED, there were 244 admissions within 3 days of an ED discharge from June 2024 to February 2025, of which 110 were manually reviewed. Of these 9 were duplicate encounters and only the most recent was used. In our sample from the overall network of EDs, there were 420,264 admissions, of which 12,419 had an ED discharge within the prior 72 hours prior. From these 90 were manually reviewed. These two sets were combined for analysis as a single set of 191 cases of 72 hour returns with admission within 3 days of an ED discharge. Within these cases, we identified 21 as having an MOD with a PPV of 11% and NNS of 9.1. Across reviewers, the median time per case review was 5 minutes (range: 3-7 min). Screening minutes per detected MOD varied by eTrigger: 27.5 for floor-to-ICU escalations, 29.0 for ED bouncebacks to ICU admission, and 45.5 minutes for the 72 hour ED returns to admission.

**Figure 1:**
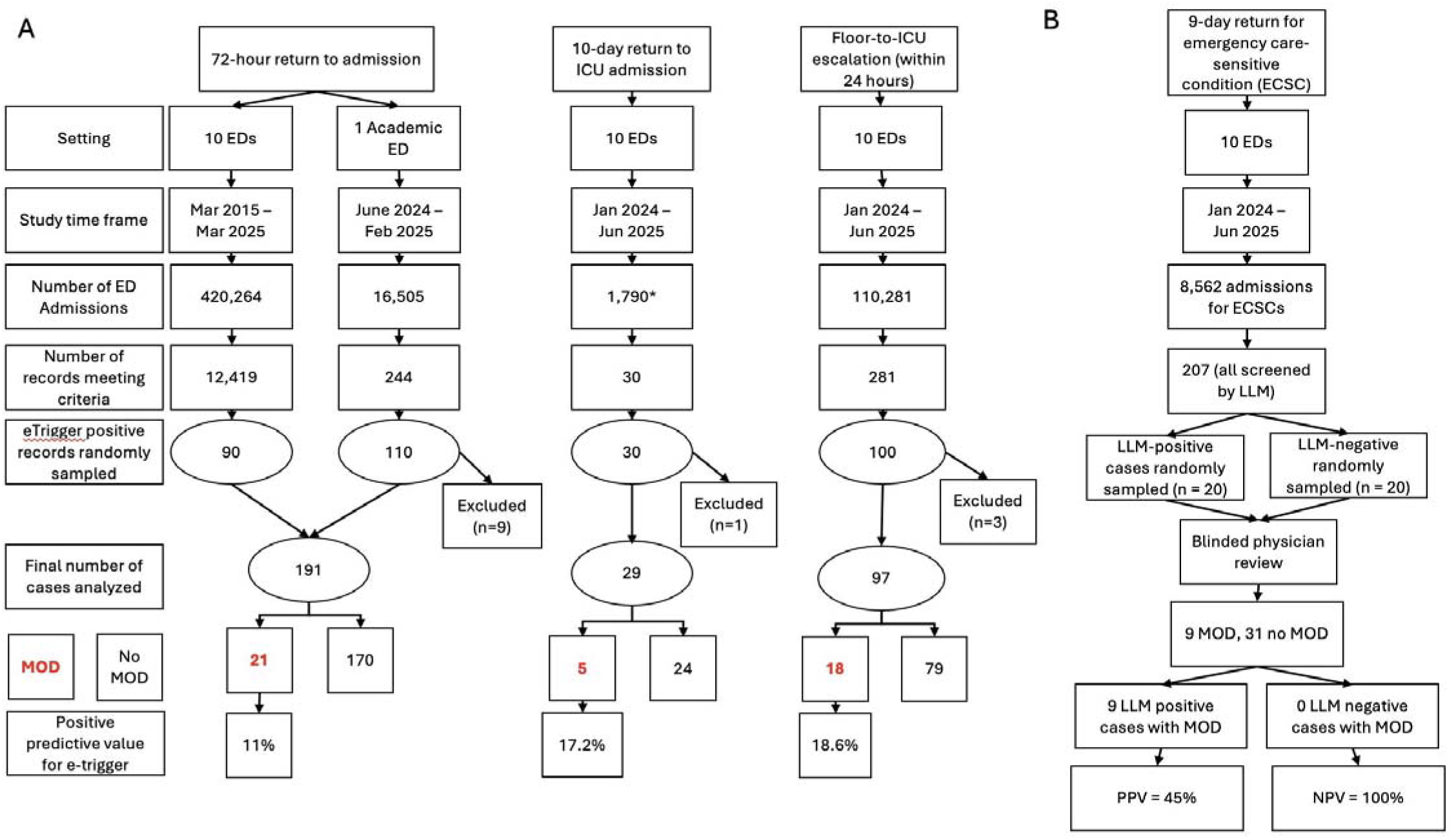
Study flow diagram for the (A) 3 72-hour return to admission trigger, 10-day return-to-admission trigger, floor admission with ICU escalation within 24 hour trigger, and (B) a hybrid, sequential eTrigger to LLM screen for 9-day emergency care sensitive conditions. * ICU admissions only

**Table 1:** Study demographics by eTrigger category from 10 hospitals (2 academic, 8 community) in the Beth Israel Lahey Health system.

|  |  | **Overall** | **10 day**<br>**return to**<br>**ICU**<br>**admission** | **72-hour return**<br>**to admission** | **9 day**<br>**emergency**<br>**condition** | **Floor admission to**<br>**ICU within 24 hours** |
| --- | --- | --- | --- | --- | --- | --- |
| **n** |  | 357 | 29 | 191 | 40 | 97 |
| **age, mean**<br>**(SD)** |  | 66.2 (17.8) | 68.9<br>(17.1) | 63.4 (19.1) | 71.0 (16.1) | 68.8 (15.4) |
| **sex, n (%)** | **Female** | 168 (47.1) | 14 (48.3) | 95 (49.7) | 19 (47.5) | 40 (41.2) |
|  | **Male** | 189 (52.9) | 15 (51.7) | 96 (50.3) | 21 (52.5) | 57 (58.8) |
| **First race,**<br>**n (%)** | **Other** | 15 (4.2) | 2 (6.9) | 10 (5.2) | 2 (5.0) | 1 (1.0) |
|  | **White or**<br>**Caucasian** | 299 (83.8) | 27 (93.1) | 149 (78.0) | 34 (85.0) | 89 (91.8) |
|  | **American**<br>**Indian** | 1 (0.3) |  | 1 (0.5) |  |  |
|  | **Asian** | 6 (1.7) |  | 3 (1.6) | 1 (2.5) | 2 (2.1) |
|  | **Black or**<br>**African**<br>**American** | 29 (8.1) |  | 23 (12.0) | 2 (5.0) | 4 (4.1) |
|  | **Hispanic** | 2 (0.6) |  | 1 (0.5) | 1 (2.5) |  |
|  | **Patient**<br>**chooses not**<br>**to disclose** | 4 (1.1) |  | 3 (1.6) |  | 1 (1.0) |
|  | **Unable to**<br>**Obtain** | 1 (0.3) |  | 1 (0.5) |  |  |
| **ethnicity,** | **Hispanic** | 22 (6.2) | 3 (10.3) | 14 (7.3) | 2 (5.0) | 3 (3.1) |
| **n (%)** | **Non-Hispanic** | 335 (93.8) | 26 (89.7) | 177 (92.7) | 38 (95.0) | 94 (96.9) |

For 72-hour returns to admission, language model screening reported 91 MODs (48.2% of all cases) with sensitivity of 85.7% (65.4-95.0%), specificity of 56.8 (49.3-64.0%), PPV of 19.8% and NPV of 97.0% (confusion matrix shown in Supplementary Table S1). For returns to ICU admission, 19 MODs were identified (65.5%), with sensitivity of 100% (56.6-100.0%), specificity 41.7% (24.5-61.2%), PPV of 26.3%, and NPV of 100%. Floor-to-ICU escalation MODs were identified in 38 (39.2%) of cases with resulting sensitivity of 55.6% (33.7- 75.4%), specificity of 64.6% (53.6-74.2%), PPV of 26.3%, and NPV of 86.4%.

There were 8,562 admissions for high risk ECSCs in the queried period (Figure 1B). Of these, 207 had one or more discharges within the preceding 9 days. The proportion of admissions with prior discharges varied by condition, ranging from 0% for aortic dissection to 10.2% for encephalitis (Table 2). Of the 20 randomly selected negative cases, blinded review identified all as true negatives (NPV = 100), while of the 20 randomly selected positive cases, blinded review identified 9 as true positives (PPV = 45%). Across all 207 ECSC encounters with prior discharges, 110 cases were identified by the LLM (53.1%) as having likely diagnostic errors on the initial visit. MOD rates varied by diagnosis ranging from 41.7% (5 of 12 cases) for spontaneous intracranial hemorrhage to 80% (4 of 5 cases) for encephalitis (Figure 2).

**Figure 2:**
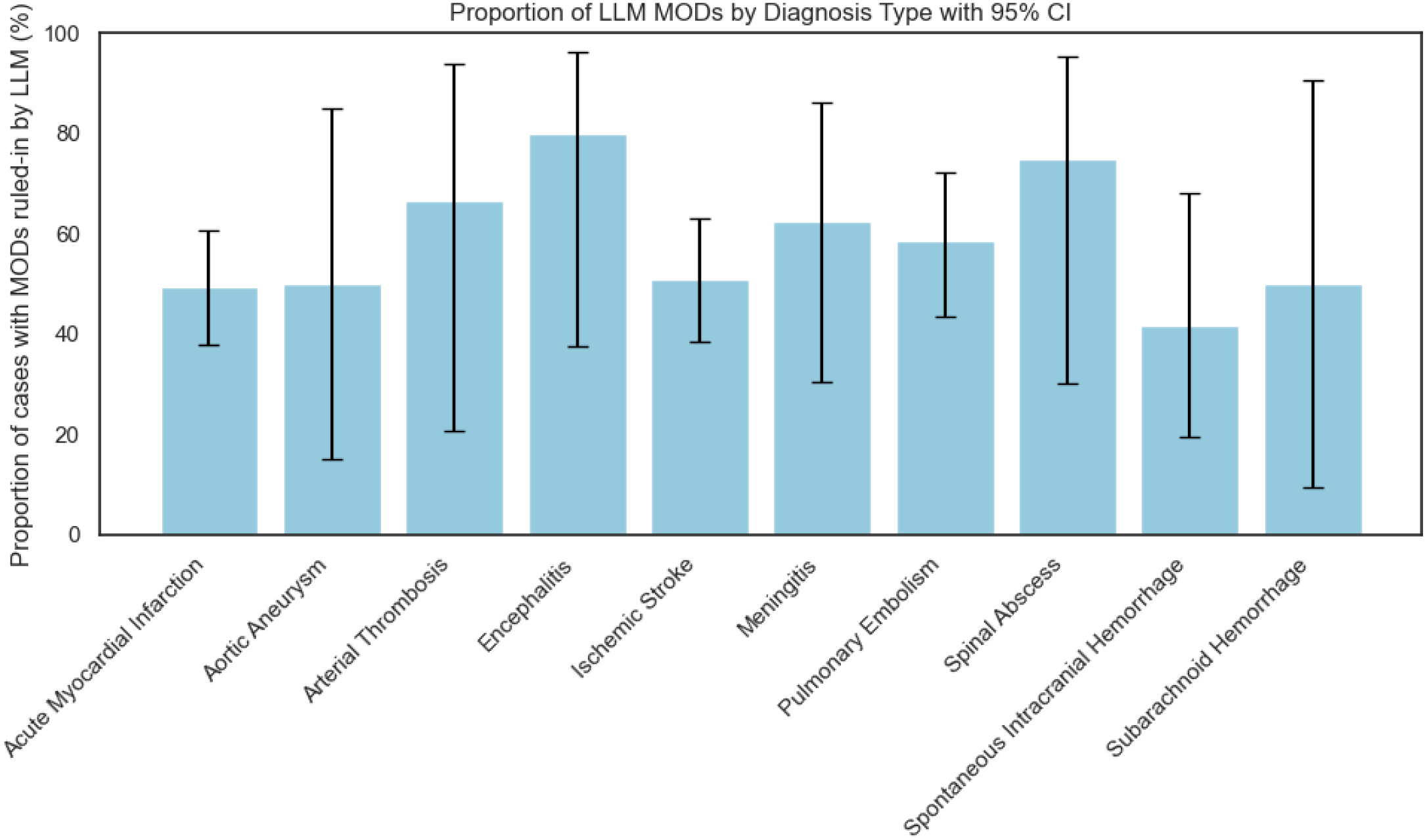
LLM screened MOD proportion by diagnosis type

**Table 2:**
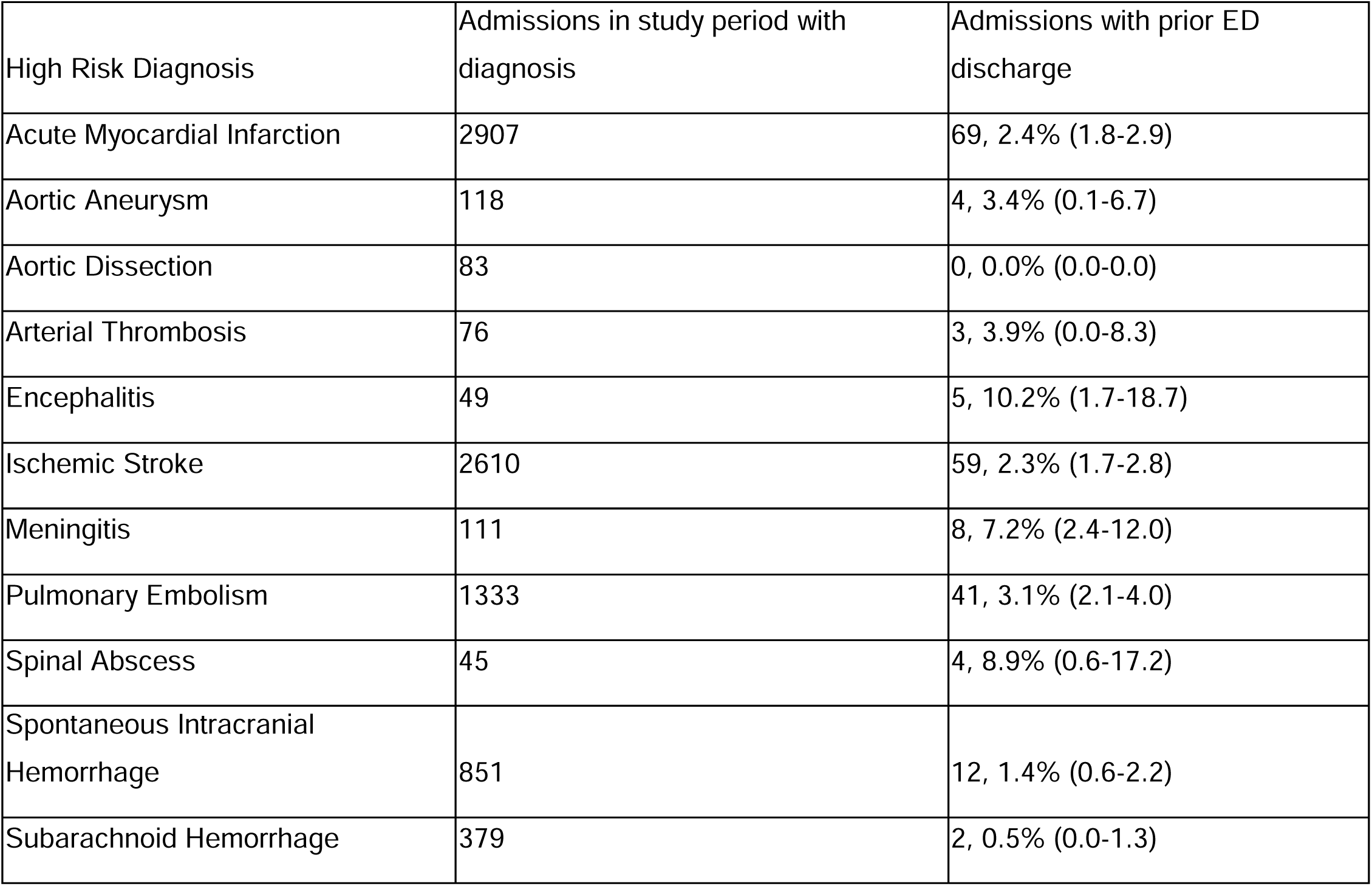
Admissions with high risk diagnoses.

Five emergency physicians from 3 hospitals participated in the quality improvement stakeholder case review, during which 14 cases were discussed (examples in Table 3). Across these 14 cases, the mean recommendation for further chart review was 4.9 (95% CI: 4.8-5.0). The systems quality implication rating was 1.4 (1.0-1.7) and the individual physician quality rating was 4.1 (3.4-4.7).

**Table 3:**
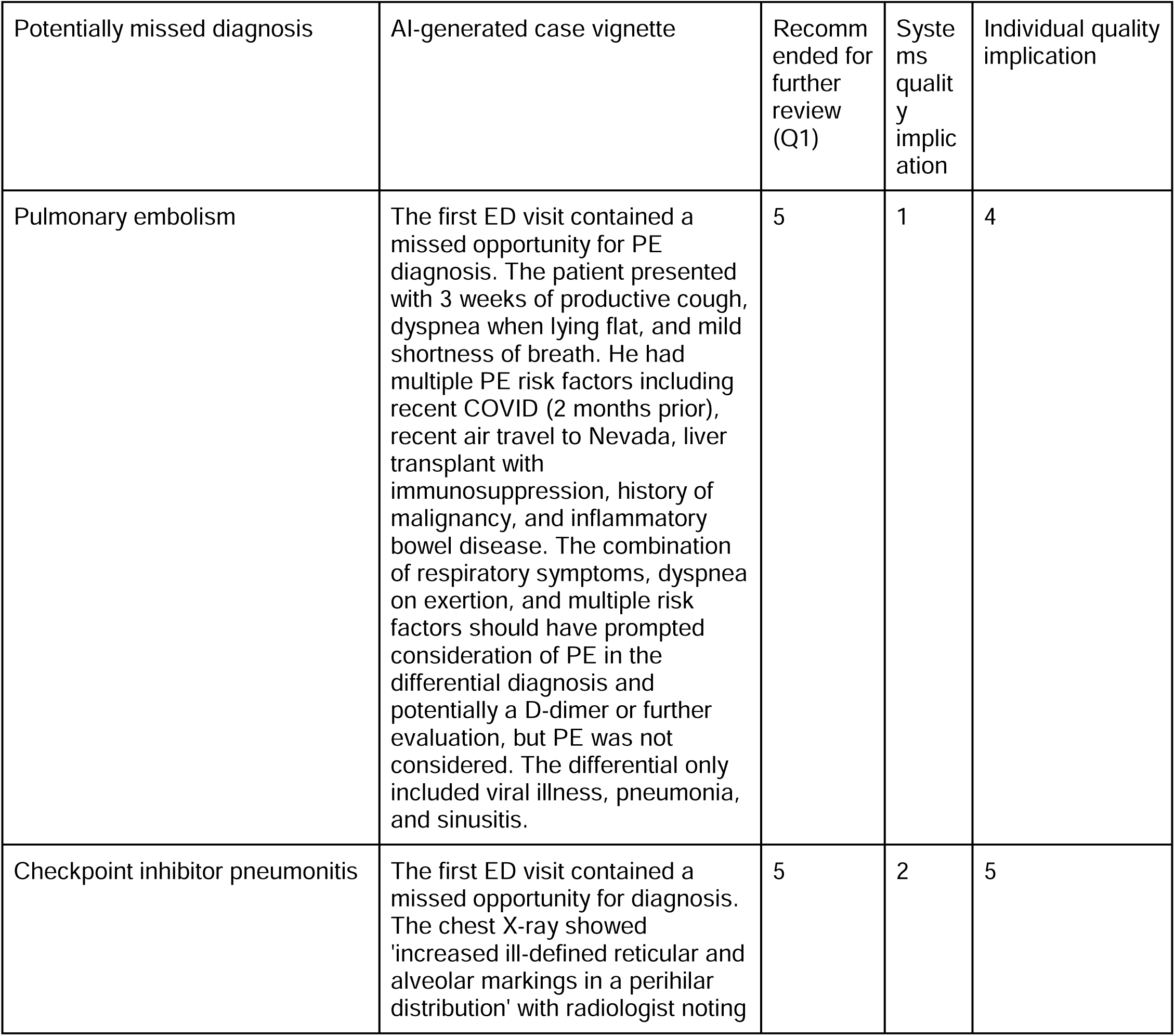
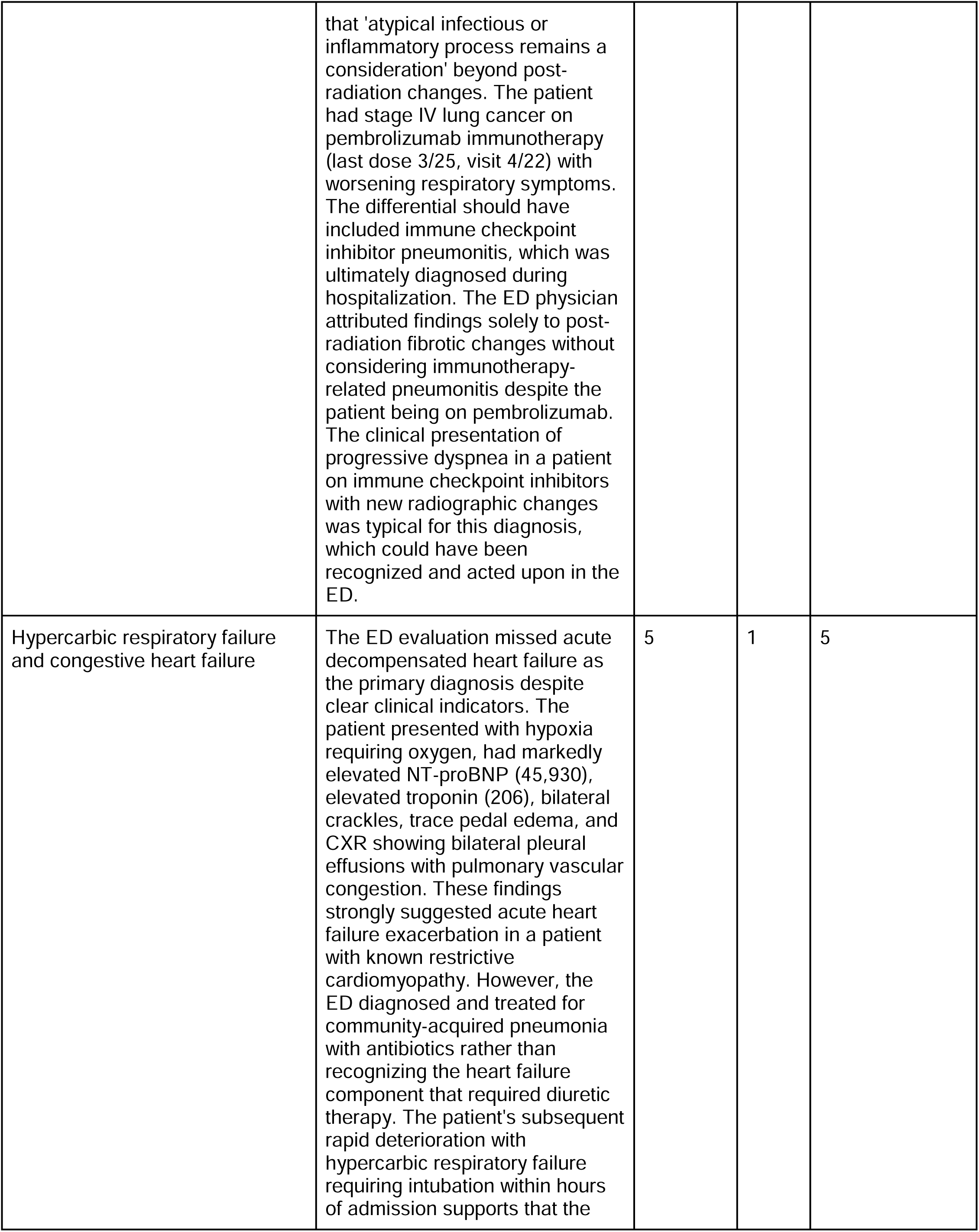
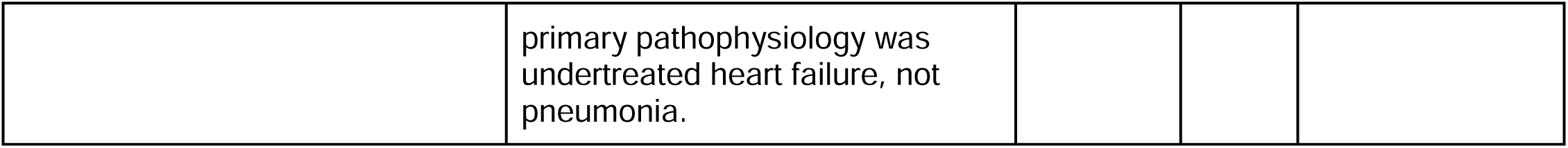
Advancing the science of emergency care using scalable MOD review. Example vignettes from ED quality and safety stakeholder meeting. The panel of emergency physician quality leaders (n = 5 drawn from 3 different hospitals with non-overlapping QA teams) were asked to reach consensus on each of these questions: Q1. Would you recommend this case for further chart review by a member of the QA team? (responses: definitely would not recommend to definitely recommend); Q2. From a systems quality perspective, is the case described by this summary potentially actionable? (responses: this case would definitely not change system quality performance to it definitely would); Q3. From an individual clinician quality perspective, is the case described by this summary potentially actionable? (responses: this case would definitely not change system quality performance to it definitely would). Consensus Likert responses for each question were recorded.

| Potentially missed diagnosis | AI-generated case vignette | Recommended for further review (Q1) | Systems quality implication | Individual quality implication |
| --- | --- | --- | --- | --- |
| Pulmonary embolism | The first ED visit contained a missed opportunity for PE diagnosis. The patient presented with 3 weeks of productive cough, dyspnea when lying flat, and mild shortness of breath. He had multiple PE risk factors including recent COVID (2 months prior), recent air travel to Nevada, liver transplant with immunosuppression, history of malignancy, and inflammatory bowel disease. The combination of respiratory symptoms, dyspnea on exertion, and multiple risk factors should have prompted consideration of PE in the differential diagnosis and potentially a D-dimer or further evaluation, but PE was not considered. The differential only included viral illness, pneumonia, and sinusitis. | 5 | 1 | 4 |
| Checkpoint inhibitor pneumonitis | The first ED visit contained a missed opportunity for diagnosis. The chest X-ray showed 'increased ill-defined reticular and alveolar markings in a perihilar distribution' with radiologist noting<br>that 'atypical infectious or inflammatory process remains a consideration' beyond post-radiation changes. The patient had stage IV lung cancer on pembrolizumab immunotherapy (last dose 3/25, visit 4/22) with worsening respiratory symptoms. The differential should have included immune checkpoint inhibitor pneumonitis, which was ultimately diagnosed during hospitalization. The ED physician attributed findings solely to post-radiation fibrotic changes without considering immunotherapy-related pneumonitis despite the patient being on pembrolizumab. The clinical presentation of progressive dyspnea in a patient on immune checkpoint inhibitors with new radiographic changes was typical for this diagnosis, which could have been recognized and acted upon in the ED. | 5 | 2 | 5 |
| Hypercarbic respiratory failure and congestive heart failure | The ED evaluation missed acute decompensated heart failure as the primary diagnosis despite clear clinical indicators. The patient presented with hypoxia requiring oxygen, had markedly elevated NT-proBNP (45,930), elevated troponin (206), bilateral crackles, trace pedal edema, and CXR showing bilateral pleural effusions with pulmonary vascular congestion. These findings strongly suggested acute heart failure exacerbation in a patient with known restrictive cardiomyopathy. However, the ED diagnosed and treated for community-acquired pneumonia with antibiotics rather than recognizing the heart failure component that required diuretic therapy. The patient's subsequent rapid deterioration with hypercarbic respiratory failure requiring intubation within hours of admission supports that the<br>primary pathophysiology was undertreated heart failure, not pneumonia. | 5 | 1 | 5 |

## Discussion

MODs are a high-priority target for ED quality and safety improvement, but systematic detection is limited by the significant effort required in case review. Here, we show that sequential screening combining eTriggers with LLMs substantially enriches for MODs compared to eTriggers alone. When compared to human reviewers, LLMs overcalled errors, but provided accurate MOD exclusion with NPVs ranging from 86.4-100% across the tested eTriggers.

In light of these findings, we propose that LLMs be used as an initial screen, either to improve the efficiency of expert adjudication of MODs or to expand error identification into new classes of patients. Our study allows for quantification of these savings. For 72-hour returns, if human reviewers only adjudicated positive charts, they would decrease effort by half while still identifying 18/21 MODs; for ICU admission within 10 days, they would cut review time by 35% with no misses. For floor to ICU within 24 hours, the NPV may not be high enough to successfully screen despite considerable time savings (61%) as only 10 of 18 MODs were identified. However, since addressing repeated errors is likely more impactful than examining outlier cases, the better yield and time savings afforded by an LLM screen may justify missing a small number of individual error cases. To test the generalizability of this approach and model how sequential screening would be implemented in the future, we developed a new integrated screen for high-risk ECSCs, resulting in a NPV of 100% and a PPV of 45%, in line with the best-performing eTriggers.

Error adjudication in the emergency department has generally focused on individual performance, rather than process improvements. Because emergency physicians operating in an episodic care capacity rarely have diagnostic follow-up on patients they see, there is significant risk for diagnostic miscalibration – i.e., misalignment between a physician’s confidence in their work up and their actual accuracy.^23^ We propose that LLM case summaries will facilitate diagnostic closure, providing individual clinicians valuable feedback to improve diagnostic calibration, even when an MOD is not present. Our quality stakeholder case review suggests that cases identified by the LLM would likely improve individual quality performance if provided to individual clinicians. Moreover, these narrative summaries in high risk patients could also be analyzed en masse to identify systems errors and formulate solutions, such as developing new pathways for communicating critical findings on final reads of CT scans that result after the patient has been discharged. Our results are hypothesis generating only; future research will need to provide individualized feedback to clinicians and meaningfully track changes in performance. However, both of these use cases reflect novel error mitigation strategies that have not previously been possible because of the high amount of expert time necessary for error identification.

Our study also suggests that LLMs may be helpful in adjudicating new classes of diagnostic error that have previously been overlooked in the MOD literature. Unlike earlier studies that used classifier models, no extensive human labeling was necessary in the development of our tool, just iterative prompt engineering in plain English using an off-the-shelf, commercially available language model. As such, this approach is scalable and translatable to new settings such as inpatient and primary care. Here, we investigated 9-day returns admissions for high-risk emergency conditions like myocardial infarction and aortic dissection. Our initial eTrigger results are in line with a recent study evaluating ECSCs using Medicare claims.^19^ However, when using sequential LLM screening, 47% of screened cases were false positives. We draw two high level conclusions from this discordance. The first is that it is very difficult to estimate incidence of MODs from administrative data; individual case screening via LLM will increase specificity. The second is that understanding the national landscape of MODs in a way that is representative of diverse populations and care settings will require new types of data consortia sharing granular EHR-level data and empowered with latest-generation LLMs.

## Limitations

Our study has several limitations. Cases came from a single network of hospitals, which may limit generalizability. We also used a single LLM—performance may vary if employing a different model. In addition, our study population demonstrated limited racial/ethnic diversity (85.5% White), which precluded assessment of fairness across demographic groups. This study also did not systematically assess the accuracy of case summaries, though authors did manually review charts of the highlighted visits that pointed to possible system or practice changes. Our study did not measure the relative severity or harm of MODs. Finally, our MOD rates are lower than the existing literature; our sequential eTrigger may have different performance with different base error rates.

## Conclusion

LLMs have excellent rule-out performance in the adjudication of MODs in a variety of eTrigger situations in the ED. Sequentially combining LLM review with human expert review outperforms other eTriggers for MOD screening, and LLM-based summaries offer new opportunities for both diagnostic excellence and identifying new systems-level error mitigation.

## Supporting information

Supplemental Figure 1 TRIPOD-LLM Checklist

## Data Availability

Data will not be shared as they include PHI.

## Supplementary Materials

## Appendix A: E-trigger criteria

1. 72 hour return to admission:

a. Inclusion Criteria

i. Patients with at least two emergency department (ED) encounters.
ii. The first (index) ED encounter must meet all of the following:

1. ED disposition is “Discharge”.
iii. The second (return) ED encounter must meet all of the following:

1. ED disposition is “Admit”.
2. A bed request was placed.
3. Arrival time is after the departure time of the first encounter.
4. The number of days between the first departure and second arrival is between 1 and 3, inclusive.
b. Exclusion Criteria

i. The second encounter has levelofcare = ‘Behavioral Health’.
ii. The department specialty of the first encounter begins with “Urgent Care”.
2. Floor-to-ICU escalation within 24 hours:

a. Inclusion criteria

i. The first (earliest) uncancelled bed request (if present) is not for Intensive Care level of care.
ii. There is a subsequent uncanceled bed request for Intensive Care level of care.
iii. The Intensive Care bed request occurs within 24 hours of ED arrival
iv. The Intensive Care bed request is the first such request for the encounter (i.e., it is the first non-canceled ICU request by request time).
b. Exclusion criteria

i. Encounters where no ICU bed request was made.
ii. Encounters where the ICU bed request occurred more than 24 hours after ED arrival.
iii. Encounters where the first uncanceled bed request was already for Intensive Care (i.e., not a transition from a lower level of care).
3. 10 day return to ICU admission:

a. Inclusion Criteria

i. Patients with at least two emergency department (ED) encounters.
ii. The first (index) ED encounter must meet all of the following:

1. ED disposition is “Discharge”.
iii. The second (return) ED encounter must meet all of the following:

1. ED disposition is “Admit”.
2. Level of care is “Intensive Care”
3. A bed request was placed.
4. Arrival time is after the departure time of the first encounter.
5. The number of days between the first departure and second arrival is between 1 and 10, inclusive.
b. Exclusion criteria

i. Encounters where the second ED visit does not result in admission to intensive care.
ii. Encounters where the second ED visit occurs on the same day as the first ED discharge, or more than 10 days after.
iii. Encounters where the second ED visit does not have a valid bed request.
4. 9 day return with high risk ECSC

a. Inclusion Criteria

i. Second “ECSC” encounter has a primary diagnosis matching one of the following categories

1. Ischemic Stroke (I63%, I64%)
2. Spontaneous Intracranial Hemorrhage (I61%, I62%)
3. Subarachnoid Hemorrhage (I60%)
4. Acute Myocardial Infarction (I21%)
5. Aortic Dissection (I71.0%)
6. Aortic Aneurysm (I71.1%–I71.9%)
7. Arterial Thrombosis (I74%)
8. Pulmonary Embolism (I26.0%, I26.9%)
9. Meningitis (G00%–G03%, A87%, A02.21%, A17.0%, A27.81%, A39.0%, A50.41%, A51.41%)
10. Encephalitis (e.g., G04%–G05%, A83%–A86%)
11. Spinal Abscess (e.g., G06.1%)
ii. The diagnosis must be documented as the primary diagnosis (isprimary = 1).
iii. There was a visit resulting in discharge between 1-9 days prior to the ECSC encounter
b. Exclusion Criteria

i. ED visits not matching the above diagnosis code or not having a diagnosis code

## Appendix B: Reviewer MOD criteria

### Reviewer Missed Opportunity for Diagnosis (MOD) criteria

For each ED encounter, was there a missed opportunity to make a correct or timely diagnosis based on the available evidence, regardless of harm? This standard is asking whether a perfect emergency physician operating at their best could have made this diagnosis, not whether we are sure we would have done so 100% of the time. For an MOD to have occurred, the patient should have signs, symptoms, laboratory or imaging findings suggestive of an alternate diagnosis that could have been made by the emergency physician.

#### Example

MOD

1. Patient with diabetes with lower extremity weakness is admitted to medicine for workup of weakness. ED labs with elevated anion gap and low bicarbonate consistent with euglycemic DKA, without specific mention or action around this diagnosis.

#### Example

not an MOD

1. Patient presents with new onset confusion and erratic behavior. Labs and CT are normal. Patient is admitted to medicine with AMS. During hospitalization, the patient is diagnosed with anti-NMDA receptor encephalitis. *Reason*: Signs and symptoms were present at time of evaluation, but this is not a diagnosis that can be made by an emergency physician as the LP results would not be available during the encounter.
2. Patient presents with back pain after lifting weights. No fevers, IV drug use, or other risk factors noted. Patient returns in 1 week with fever and back pain, found to have spinal epidural abscess. *Reason:* The signs and symptoms at time of evaluation were not suggestive of SEA as the diagnosis.

The following factors are associated with the presence of a missed opportunity for diagnosis (ref, see graphic below).

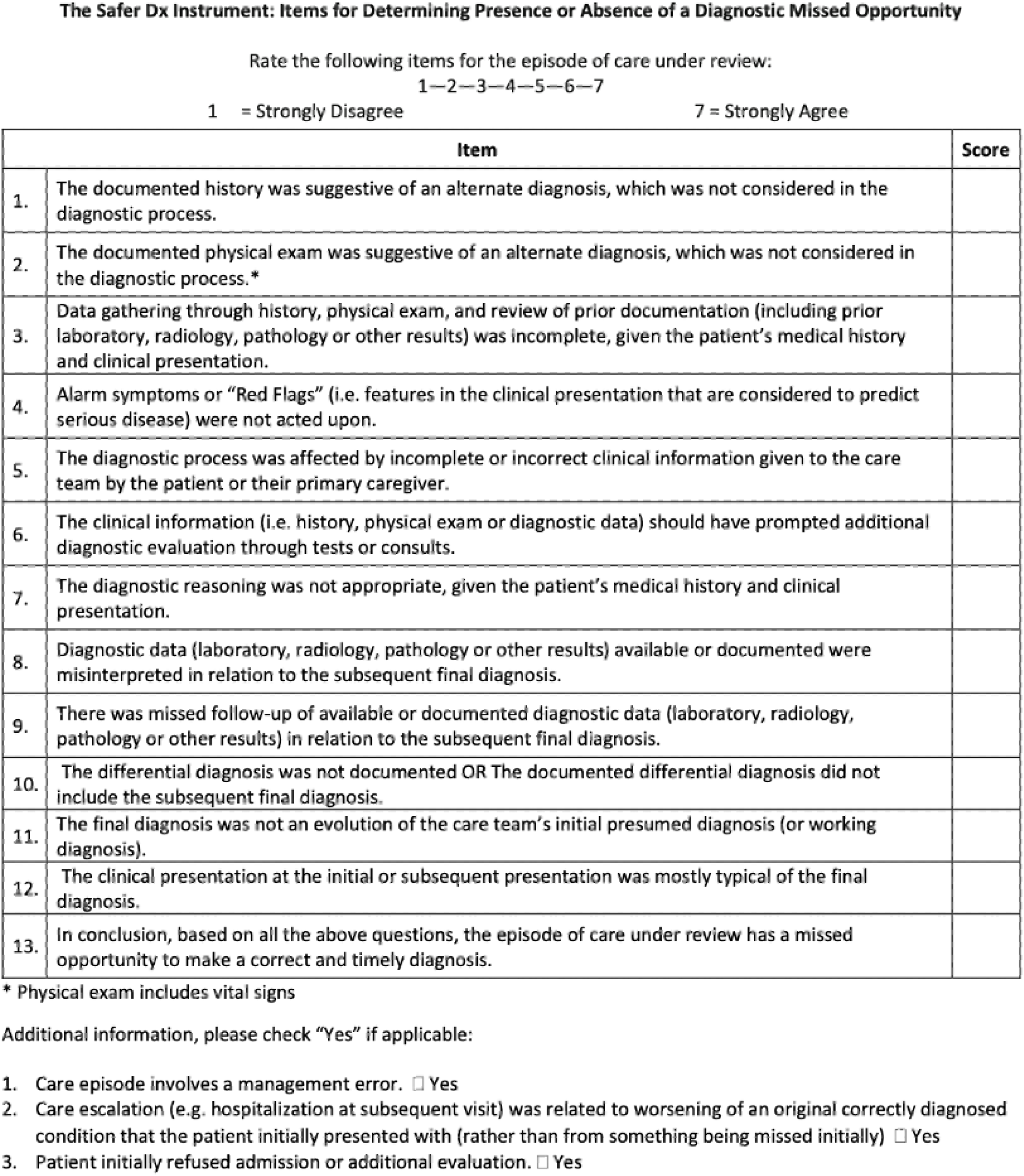

## Appendix C: LLM MOD review prompt

**You are ED-Diagnostic-Error-Review-Bot, reasoning like an expert emergency physician.**

**Task: do *all* of the following:**

1. **Read every data block once, think, then decide** whether the first ED visit contained a *missed opportunity for diagnosis* (MOD) considering

- the 13 items of the **SaferDx Instrument**
- the MOD definition & examples given below (*built-in for your reference—do not quote back*).
2. Provide a likelihood estimate (0-100%) that there was a missed opportunity for diagnosis. You are NOT to consider management, only diagnosis.
3. Make a **Yes/No call**:

- **Yes** if your clinical judgment clearly supports a missed opportunity.
- **No** otherwise.
4. List up to 5 details that anchor your determination
5. Output a JSON and nothing else:

~~~
'''
{{
"mod_decision": "Yes", // or "No" "mod_likelihood_estimate": "0-100%"
"mod_rationale": "Brief narrative summary of why MOD is or is not present, focused on diagnostic reasoning."
}}
'''
~~~

Here are the SaferDx criteria to consider:

1. The documented history was suggestive of an alternate diagnosis, which was not considered in the diagnostic process.
2. The documented physical exam was suggestive of an alternate diagnosis, which was not considered in the diagnostic process.
3. Data gathering through history, physical exam, and review of prior documentation (including prior laboratory, radiology, pathology, or other results) was incomplete, given the patient’s medical history and clinical presentation.
4. Alarm symptoms or “red flags” (i.e., features in the clinical presentation that are considered to predict serious disease) were not acted upon.
5. The diagnostic process was affected by incomplete or incorrect clinical information given to the care team by the patient or their primary caregiver.
6. The clinical information (i.e., history, physical exam or diagnostic data) should have prompted additional diagnostic evaluation through tests or consults.
7. The diagnostic reasoning was not appropriate, given the patient’s medical history and clinical presentation.
8. Diagnostic data (laboratory, radiology, pathology or other results) were available or documented were misinterpreted in relation to the subsequent final diagnosis.
9. There was missed follow-up of available or documented diagnostic data (laboratory, radiology, pathology or other results) in relation to the subsequent final diagnosis.
10. The differential diagnosis was not documented OR the documented differential diagnosis did not include the subsequent final diagnosis.
11. The final diagnosis was not an evolution of the care team’s initial presumed diagnosis (or working diagnosis).
12. The clinical presentation at the initial or subsequent presentation was mostly typical of the final diagnosis or more amenable to a correct and timely diagnosis.
13. In conclusion, based on all the above questions, the episode of care under review has a missed opportunity to make a correct and timely diagnosis.

---

### Examples

MOD:

Patient with diabetes with lower extremity weakness is admitted to medicine for workup of weakness. ED labs with elevated anion gap and low bicarbonate consistent with euglycemic DKA, without specific mention or action around this diagnosis.

Not an MOD:

Patient presents with new onset confusion and erratic behavior. Labs and CT are normal. Patient is admitted to medicine with AMS. During hospitalization, the patient is diagnosed with anti-NMDA receptor encephalitis.

Reason: Signs and symptoms were present at time of evaluation, but this is not a diagnosis that can be made by emergency physician as the LP results would not be available during the encounter.

Patient presents with back pain after lifting weights. No fevers, IV drug use, or other risk factors noted. Patient returns in 1 week with fever and back pain, found to have spinal epidural abscess. Reason: The signs and symptoms at time of evaluation were not suggestive of SEA as the diagnosis.

~~~
---
### Case Documentation
{case_details}
---
### Output JSON:
~~~

## Appendix D: Included clinical documentation for each of the eTrigger prompts

72 hour return to admission, 10 day return to ICU admission, 9 day return to ECSC high risk condition:

- Encounter 1:

- ED physician notes
- ED nursing flowsheets (vital signs, nursing documentation)
- Non-ED documentation (e.g., case management notes, consults)
- ED labs - all labs resulting prior to patient departure
- ED imaging reports as resulted prior to patient departure
- Encounter 2:

- ED physician notes
- ED nursing flowsheets (vital signs, nursing documentation)
- Non-physician documentation (e.g., case management notes, consults)
- Notes from first 48 hours of hospitalization with exclusions*

Floor admission with ICU escalation within 24 hours:

- ED physician notes
- ED nursing flowsheets (vital signs, nursing documentation)
- Non-ED documentation (e.g., case management notes, consults) within 48 hours with exclusions*
- ED labs - all labs resulting prior to patient departure
- ED imaging reports as resulted prior to patient departure

* Excluded note types:

ACP (Advance Care Planning)
Brief Op Note
Care Coordination Note
Code Documentation
Code Status and Healthcare Proxy
Diabetic Education
Discharge Instructions
Discharge Instr – Activity
Discharge Instr - AVS First Page
Discharge Instr - Diet
Discharge Instr – Lab
Discharge Instr - Other Orders
Discharge Instr - Pt Ed Handouts
Discharge Instr-Facility, Services & DME
Discharge Instr – Wound
Downtime Event Note
ED AVS Snapshot
ED Information Exchange
ED Procedure Note
Global ED Visit
H&P (View-Only)
Interval H&P Note
IP AVS Snapshot
IV Therapy Notes
Liaison Communication
Letter
Medical Student
Moderate Sedation
NCDR Registry
OR Nursing
Op Note
Patient Care Conference
PCU Note
Research
Treatment Plan
Utilization Review
Wound Ostomy Note

## Appendix E: LLM MOD review for high risk ECSCs

**You are ED-Diagnostic-Error-Review-Bot, reasoning like an expert emergency physician.**

**Task: do *all* of the following:**

1. Read every data block once, think, and then decide about the following:
2. Was the {CRITICAL_DIAGNOSIS_TYPE} present at the second ED encounter or did it develop during hospitalization? Answer: Before or in ED|After admission
3. Did the first ED visit contained a *missed opportunity for diagnosis* (MOD) related to {CRITICAL_DIAGNOSIS_TYPE}?

- the 13 items of the **SaferDx Instrument**
- the MOD definition & examples given below (*built-in for your reference—do not quote back*).
4. Provide a likelihood estimate (0-100%) that there was a missed opportunity for diagnosis. You are NOT to consider management, only diagnosis.
5. Make a **Yes/No call**:

- **Yes** if your clinical judgment clearly supports a missed opportunity.
- **No** otherwise.
6. List up to 5 details that anchor your determination
7. Output a JSON and nothing else:

~~~
'''
{{
"present_in_ED": "<Before or in ED|After admission>",
"present_in_ED_evidence": "<Brief narrative explaining in what phase of the patient’s care the stroke became apparent>",
"mod_decision": "<Yes|No>",
"mod_likelihood_estimate": "<0-100%>",
"mod_rationale": "<BRIEF narrative summary of why the diagnosis was considered missed or not>"
}} ‘’’
… Remainder of prompt is the same as Appendix C…
~~~

---

**Supplementary Table 1:**
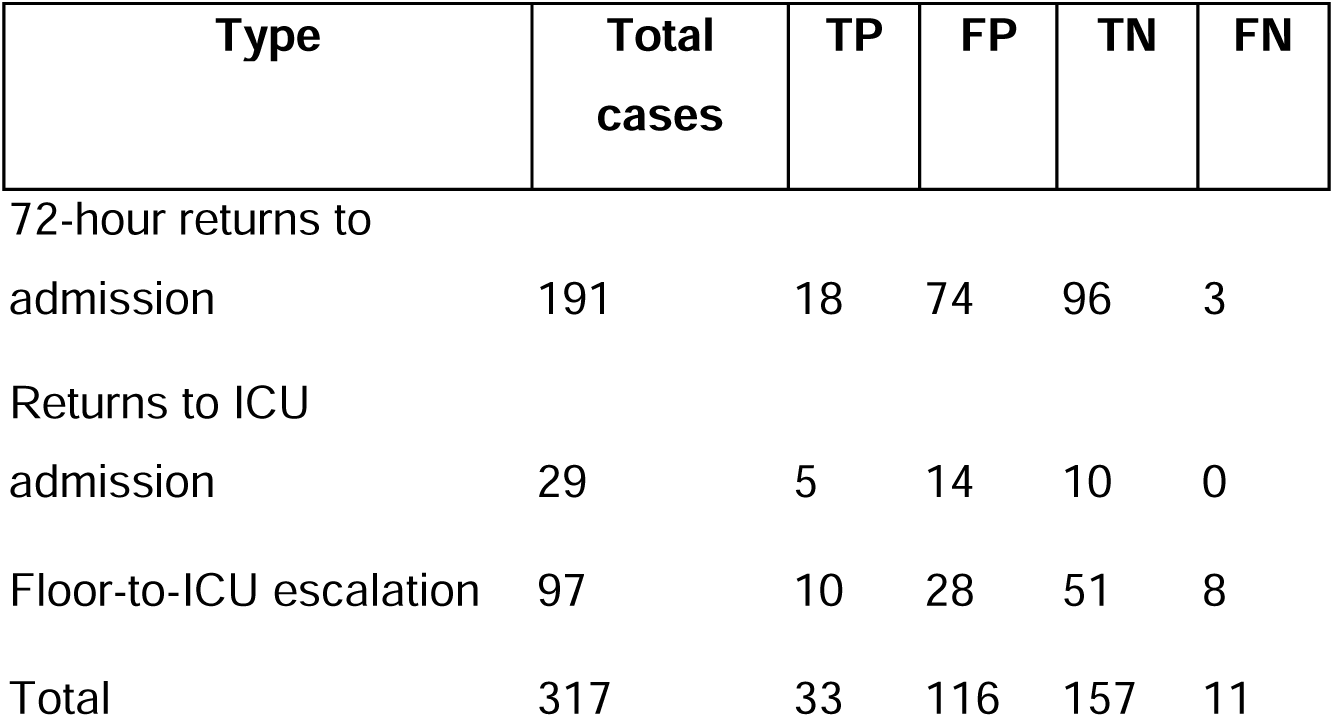
LLM screening confusion matrix.

| Type | Total cases | TP | FP | TN | FN |
| --- | --- | --- | --- | --- | --- |
| 72-hour returns to admission | 191 | 18 | 74 | 96 | 3 |
| Returns to ICU admission | 29 | 5 | 14 | 10 | 0 |
| Floor-to-ICU escalation | 97 | 10 | 28 | 51 | 8 |
| Total | 317 | 33 | 116 | 157 | 11 |

## Data Sharing Statement

Data will not be shared as they include PHI.

## Acknowledgements

This research received no specific grant from any funding agency in the public, commercial, or not-for-profit sectors.

